# Individual Differences in Effects of Stressful Life Events on Childhood ADHD: Genetic, Neural, and Familial Contributions

**DOI:** 10.1101/2025.03.18.25324229

**Authors:** Seung Yun Choi, Jinwoo Yi, Junghoon Park, Eunji Lee, Bo-Gyeom Kim, Gakyung Kim, Yoonjung Yoonie Joo, Jiook Cha

**Author notes:** **Corresponding author** Jiook Cha, PhD Department of Psychology, Seoul National University, Gwanak-ro 1, Gwanak-gu, Seoul, 08826, Republic of Korea.

## Abstract

**Background:** This study elucidates the intricate relationship between stressful life events and the development of ADHD symptoms in children, acknowledging the considerable variability in individual responses. By examining these differences, we aim to uncover the unique combinations of factors contributing to varying levels of vulnerability and resilience among children.

**Methods:** Utilizing longitudinal data from the Adolescent Brain Cognitive Development study (baseline: N=6303, age=9.9), we applied Generalized Random Forest to model the non-linear relationships among genetic predispositions, brain features, and environmental factors.

**Results:** Significant individual variability was observed in children’s ADHD symptoms post-stress, particularly at the 1-year and 2-year follow-ups. At the 1-year follow-up, increased vulnerability was indicated by heightened parental mental health problems and a lower polygenic risk score for smoking. By the 2-year follow-up, escalated parental mental health disorders, higher ADHD polygenic risk scores, and altered structural connectivity in the cognitive control network were significant contributors to individual differences.

**Conclusions:** These findings underscore the importance of integrating environmental, genetic, and neural variables to identify children vulnerable or resilient to developing ADHD symptoms following early-life stress. This study demonstrates how multimodal data combined with non-parametric machine-learning can advance precision psychology and psychiatry, aiding targeted support for affected children.

## Introduction

The nexus between early-life stress and an elevated risk for developing ADHD has been consistently documented (Greeson et al., 2014; Humphreys et al., 2015, 2019). Despite this established connection, the question of why only certain children develop ADHD in response to early-life stress while others remain resilient still persists. The interplay of neural, genetic, and environmental factors during childhood—a critical period of significant brain development—may account for this inter-subject variability in ADHD outcomes (Bouchard Jr. & McGue, 2003; Gilmore, Knickmeyer, & Gao, 2018; Halperin & Healey, 2011; Marsh, Gerber, & Peterson, 2008; Parasuraman & Jiang, 2012). Research with mouse models has shown that early-life stress can lead to enduring changes in dopaminergic neurons through glucocorticoid-mediated epigenetic regulation, particularly when combined with specific genetic vulnerabilities (Niwa et al., 2013). These modified dopaminergic pathways, integral to brain functions such as cognitive control and motivation, may be also implicated in ADHD pathophysiology (Cools, 2016; Swanson et al., 2007). Thus, understanding this dynamic interplay is essential for comprehending the differences in how children respond to similar stressful events, with not all developing ADHD symptoms.

Literature often highlights isolated aspects of this interplay. For instance, some studies focus solely on neural influences, such as decreased cortical thickness in temporal and parietal regions, which mediate the relationship between social deprivation and increased ADHD symptoms in children (McLaughlin et al., 2014). Specific brain areas, including the temporal gyrus and internal capsule, are also linked to ADHD symptoms following stressful experiences in children (Humphreys et al., 2019). However, the influence of these neural variables on ADHD symptomatology may interact with genetic and environmental factors (Cortese, 2012; Yadav et al., 2021).

Genetic factors, often represented by polygenic risk scores (PRS), may play a crucial role in determining an individual’s susceptibility to ADHD symptoms following stressful experiences by interacting with brain structure or function. ADHD PRS is associated with ADHD in children, influencing brain functional connectivity (i.e., caudate-parietal cortex, nucleus accumbens-occipital area) (Hermosillo et al., 2020). Total cortical gray matter volume mediates the relationship between PRS for ADHD comorbid with disruptive behavior disorder and externalizing problems in late childhood (Teeuw et al., 2023). Additionally, ADHD PRS predicts the mean fractional anisotropy (FA) of white matter regions, with this FA being significantly linked to ADHD symptom scores (Albaugh et al., 2019). These findings suggest that children’s genetic predisposition may shape the brain’s response to early-life stress, thereby influencing the ADHD development.

Environmental factors also contribute to children’s ADHD symptoms following stress exposure by interacting with genetic and neural factors. Family dynamics, interpersonal support, and neighborhood deprivation show significant association with ADHD symptoms in children, while these environmental stressors are also linked to gray matter volume and cortical thickness (Jeong et al., 2023). Children in more chaotic households tend to exhibit more ADHD symptoms, and their ADHD PRS significantly contributed to these chaotic environments, indicating a gene-environment interaction in ADHD development (Agnew-Blais et al., 2022).

Examining the interplay of genetic predispositions, neural, and environmental factors in relation to early-life stress and ADHD development demands an integrated approach. Overlooking this interplay may result in a skewed interpretation. We address this by employing Generalized Random Forest (GRF), a non-parametric machine-learning method adept at modeling the complex and non-linear relationships among these factors. GRF extends the traditional random forest algorithm by integrating parametric causal inference with statistical examination of potential confounders (Athey, Tibshirani, & Wager, 2019). The flexible and inductive approach enhances the detection of subtle heterogeneity patterns not feasible with conventional methods where researchers preselect predictors and examine interaction effects by testing each variable individually (Shiba et al., 2021). The efficacy of GRF is supported across various studies: natural disaster on cognitive disability and mental health (Shiba et al., 2021, 2022), social disconnection on suicidality (Solomonov et al., 2023), and efficacy of therapeutic interventions (Goligher et al., 2023).

We aim to elucidate how stressful life events uniquely influence the development of ADHD symptoms in children, considering the multivariate roles of neural, genetic, and environmental factors. By identifying patterns within these multimodal factors, we seek to uncover the varying degrees of vulnerability and resilience among children exposed to stress. These insights could help researchers and clinicians identify subpopulations needing targeted interventions. Ultimately, understanding these risk factors could lead to the development of strategies designed to protect children from developing ADHD after experiencing stress.

## Methods

### Participants

A total of 11,878 participants aged 9–10 were drawn from the Adolescent Brain Cognitive Development (ABCD) study, a prospective longitudinal cohort study conducted across 21 sites in the United States (Data Release 4.0; http://abcdstudy.org) (Casey et al., 2018). Since our study examined the association of stressful life events and ADHD symptoms in a longitudinal design (i.e., baseline, 1-year, and 2-year follow-ups), participants were excluded if they missed stressful events data at baseline; missed ADHD symptom data at each time point; or reported non-European ancestry. To ensure consistent ancestry, we included only individuals with European ancestry, which comprised almost 80% of the sample (N=6,555). We did not impute missing variables because most missing data were from non-European ancestry, and the variables related to stressful events and ADHD symptoms were crucial for our analyses. Therefore, we opted for conservative analyses. The final sample size was 6303 at baseline (male=53.2%, *M*age=9.9), 5664 at 1-year follow-up (male=53.3%), and 4641 at 2-year follow-up (male=53.6%) (Figure 1).

**Figure 1.**
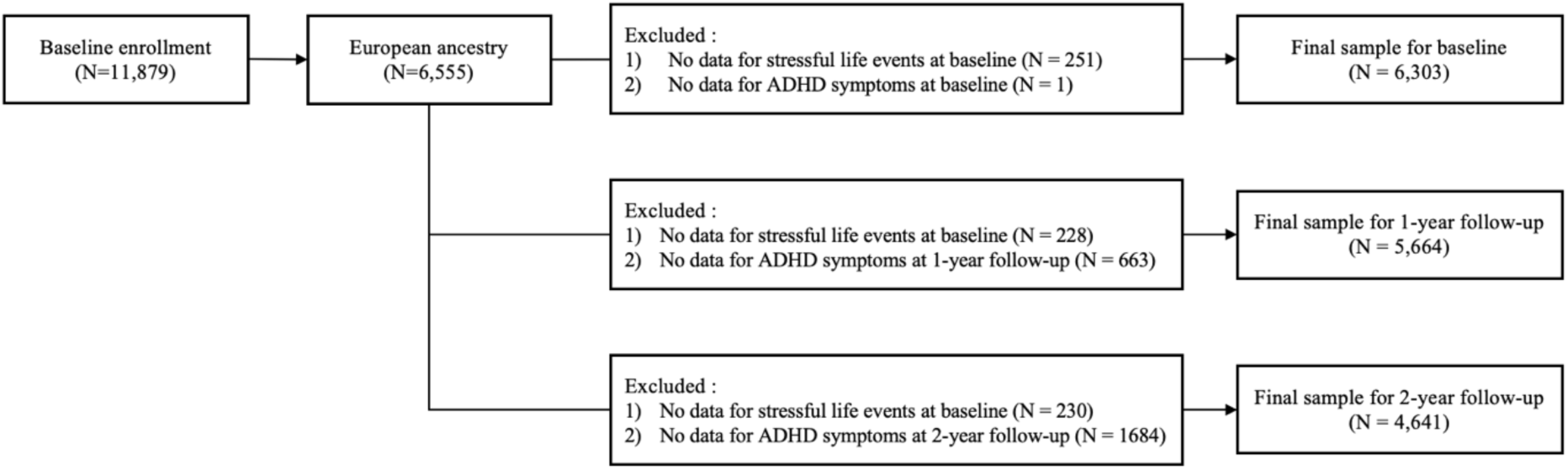
Flow chart of study sample selection.

### Ethical considerations

All parents provided written informed consent and children provided assent to the research protocol approved by the institutional review board at each of the 21 sites.

## Measures

### Stressful life events

The stressful life events were measured by the Kiddie Schedule for Affective Disorders and Schizophrenia – post-traumatic stress disorder (KSADS-PTSD) reported by the parents (Kaufman et al., 1997). Out of the 13 events in the questionnaire, seven stressful events (i.e., experienced or witnessed terrorism, death or mass destruction in a war zone, community violence, severe car accident, fire, natural disaster, or other significant accidents that require medical attention or treatment) were selected. The item was scored as 1 if the child experienced at least one event, and 0 if the child did not experience any events.

### ADHD

Children’s ADHD symptoms were assessed using a parent-reported DSM-oriented Childhood Behavior Check List (CBCL) (Achenbach & Edelbrock, 1991) at baseline, 1-year, and 2-year follow-ups. The parents answered items on a three-point Likert scale, with higher values indicating more ADHD symptoms. The reliability and validity of the CBCL was shown by prior studies (Biederman et al., 2001, 2021).

### Covariates

We included sociodemographic, genetic, and brain features from the baseline (Figure 2), assuming the potential role of these factors as confounders in the association between stressful events and ADHD symptoms.

**Figure 2.**
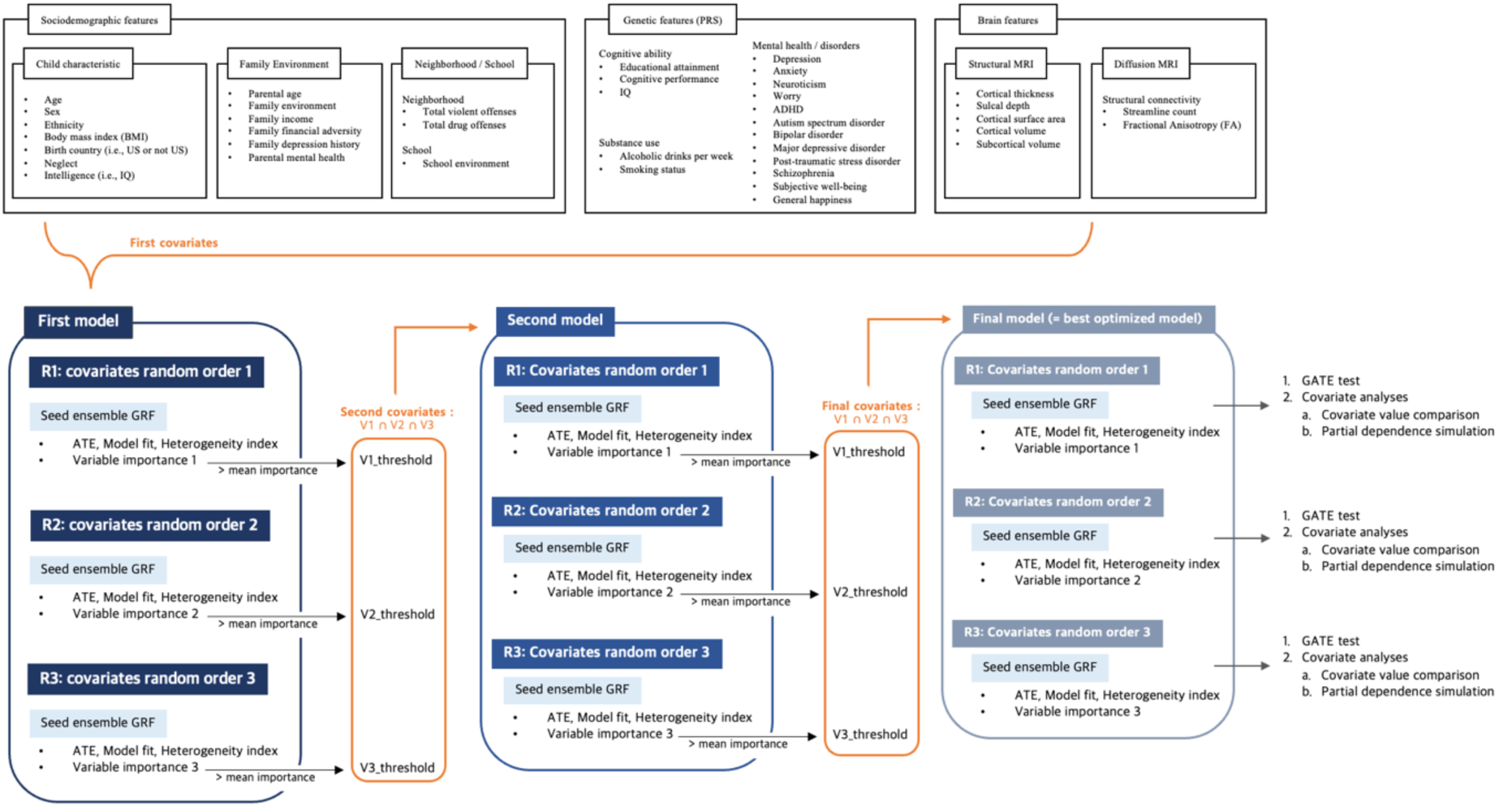
Framework of three-step analysis in Generalized Random Forest (GRF) analysis. The three-step analysis involved fitting the model with different covariate orders and seed ensembles to enhance robustness of the results (i.e., Random Iteration 1, 2, and 3). To find the optimal model, we progressively reduced the model size by retaining only the intersection of covariates that were above the mean importance in all three iterations. This optimization process narrowed down the covariates to those significantly contributing to individual differences, resulting in the final model. We then applied the GATE test to the final model to validate the existence of individual difference and conducted covariate analysis to identify potential risk and protective factors. ATE = average treatment effect; PRS = polygenic risk score; BMI = body mass index; R1 = random iteration 1; R2 = random iteration 2; R3 = random iteration 3; V1 = variable importance 1; V2 = variable importance 2; V3 = variable importance 3; GATE = group average treatment effect.

The sociodemographic characteristics consist of the child’s characteristics (i.e., age, sex, ethnicity, BMI, birth country, abuse, neglect, and IQ) and environment of family, neighborhood, and school. Family environment included parental age, marital status, parental education level, income, family conflict, family financial adversity, family depression history, and parental mental health (e.g., externalizing problems, depressive problems, etc). Neighborhood environment included rates of total violent and drug offenses in the neighborhood (United States Department Of Justice. Office Of Justice Programs. Federal Bureau Of Investigation, 2012). School environment evaluated the child’s perception of school climate. See Appendix S1 for detailed description of measurements.

For genetic characterization of the participants, we utilize polygenic risk scores (PRS) of 17 complex traits as genetic features: educational attainment, cognitive performance, IQ, depression, anxiety, neuroticism, worry, ADHD, autism spectrum disorder, bipolar disorder, major depressive disorder, PTSD, schizophrenia, subjective well-being, general happiness, alcoholic drinks per week, and smoking status. These PRSs were derived using publicly accessible genome-wide association studies (GWAS) summary statistics. See Appendix S1 and Table S1 for specific GWAS datasets, quality control measures, population stratification adjustment, and the methodology employed for polygenic scoring.

The brain morphometric features (i.e., surface area, thickness, sulcal depth, and volume) and diffusion measures (i.e., FA and streamline count) were included in the analyses. T1-weighted 3D structural images (1-mm voxel resolution) were acquired using either a 3T Siemens, Phillips, or General Electric scanner with a 32-channel head coil. We applied ABCD quality control conditions to exclude low quality brain images. Cortical and subcortical estimates were extracted using FreeSurfer 7.1.1 (https://surfer.nmr.mgh.harvard.edu). Cortical regions were labeled with Desikan-Killiany atlas (Desikan et al., 2006) and subcortical regions were segmented using automatic brain segmentation tool. Further details on image acquisition and preprocessing can be found in elsewhere (Casey et al., 2018; Hagler Jr et al., 2019).

We used diffusion spectrum images, which had undergone quality control according to a protocol (Hagler Jr et al., 2019) by the ABCD Data Analysis and Informatics Center. With the quality controlled data, we performed the preprocessing procedure using MrTrix3 (Tournier et al., 2019) and FSL (Jenkinson, Beckmann, Behrens, Woolrich, & Smith, 2012). To analyze brain structural connectivity, we generated 84×84 whole-brain individual connectome using MRtrix3 software. See Appendix S1 for detailed descriptions of diffusion image preprocessing steps.

### Generalized Random Forest and Statistical Analysis

We utilized GRF to examine the environmental, neural, and genetic variables underlying the individual differences in children’s ADHD symptoms following stress exposure. Traditional mediation and moderation analyses, while effective for testing associations between treatment and outcome with a single variable, often fall short in unraveling multivariate interactions. The non-parametric machine-learning approaches like GRF offer a more nuanced understanding of these sophisticated dynamics by modeling non-linear, multivariate relationships. This capability is particularly beneficial in our context where multiple factors are intricately intertwined. Therefore, we opted for GRF to capture these interactions more effectively.

GRF ensures reliable estimation via training trees on one data subset and assessing treatment effects on a separate subset (Wager & Athey, 2018). It repeatedly splits the data, creating decision trees by maximizing treatment effect differences (Wiemken et al., 2021). This approach mitigates overfitting and enhances treatment effect reliability. Additionally, GRF reduces confounding bias by applying doubly robust estimator (i.e., augmented inverse-propensity weighting) when estimating treatment effect (Glynn & Quinn, 2010).

GRF estimates both the average treatment effect (ATE) and individual treatment effect (ITE) on outcome. ATE measures the difference in the mean level of ADHD symptoms that would have been with and without the treatment (i.e., stressful events) (Shiba et al., 2021). A significant ATE indicates that, on average, stress-exposed children exhibit more ADHD symptoms than non-exposed children. ITE, on the other hand, provides information about individual-level effects, highlighting individual differences. Calibration tests evaluate estimate validity, with model fit and heterogeneity indices close to 1 indicating good calibration and accurate treatment effect estimation (Athey & Wager, 2019).

### Data preparation

Before training GRF models, we conducted regression analysis to identify brain features significantly associated with ADHD symptoms at each time point, controlling for sociodemographic factors and PRSs. This step reduces data dimensionality and focuses on the most relevant features. We found 29 brain features significantly related to ADHD at baseline and three at the 1-year and 2-year follow-ups (p FDR < .05; Table S2). Including sociodemographic and genetic variables, we had a total of 71 covariates at baseline, 55 covariates at 1-year and 2-year follow-ups. The abuse variable was omitted from the analyses due to its near-zero variance (p > .05), indicating redundancy.

### Analyses

Incorporating these covariates, we constructed GRF models to analyze the ATE and ITE of stressful events on ADHD symptoms at each time point. Still, due to the machine-learning nature of GRF, treatment effects can vary based on seed number and covariate order. The large number of covariates may also weaken the estimation power (Athey & Wager, 2019), necessitating model optimization. Therefore, we conducted a three-step analysis approach (i.e., first, second, and final model) (Figure 2).

The three-step analysis enhances the reproducibility of results by (1) randomly changing the order of covariates three times, resulting in Random Iteration 1, 2, and 3, and then (2) using a seed ensemble technique. Instead of relying on one seed, the seed ensemble technique combines three forests built with different random seeds to make a big forest, improving result reliability.

Furthermore, the three-step analysis helps identify the optimal model through feature (covariate) selection based on variable importance. The variable importance provides the estimate and ranking of variables based on their splitting frequency (Wang & Yang, 2022). Specifically, in the ‘*first model’*, we used all identified covariates when fitting the model. After fitting each model with different covariate orders and seed ensembles (i.e., Random Iteration 1, 2, and 3), only covariates above the mean importance estimate were retained. The intersection of these covariates across iterations formed the second covariate set. Using this covariate set, the ‘*second model’* was fitted following the same procedure, resulting in a sparser set of final covariates. The ‘*final model’* was then fitted with these covariates through repeated procedures (Figure 2). This optimization process narrows down the covariates to those that significantly contribute to individual differences, yielding three final models (i.e., Random Iteration 1, 2, and 3). We compared the results of three iterations to check the robustness. The significance and calibration of the models were assessed using ATE estimate and model calibration test.

Based on our final model, we validated individual differences using the group average treatment effect (GATE) test (Athey & Wager, 2019). The GATE test stratifies the study sample into groups based on the predicted value of ITEs and compares the estimated ATE of these groups using a t-test. We ranked the participants into three groups: low, middle, and high-risk group. A significant difference between groups confirms individual differences. A monotonic increase in estimated ATE across these groups is also critical as it confirms that the treatment effect is lowest in the low-risk group and highest in the high-risk group.

As a final step, we applied our final model to compare covariate values between low and high-risk groups for ADHD symptoms, aiming to detect potential risk and protective factors. While this test effectively highlights group differences, its univariate nature limits its capacity to address multivariate interactions among covariates. To overcome this, we also performed partial dependence simulations for the covariates (Friedman, 2001; Yi et al., 2024). For each covariate, we generated 100 synthetic samples, each representing various percentile values of that covariate while keeping other covariates fixed at their median values. These samples were input into the final model to observe how the simulated individual treatment effects varied with the covariate percentiles. An increase in treatment effect suggests that a covariate is likely a risk factor for the impact of stressful events on children’s ADHD symptoms, while a decrease suggests a protective factor. This approach offers insights into how each covariate influences ADHD symptoms considering multivariate interactions.

We used the R package ‘*grf ’* (v.2.1.0). All continuous variables were standardized (z-scaled) prior to analysis. Also, since the ABCD data is recruited from 21 sites across the U.S., we considered the site as a clustering factor to adjust its influence on the outcome measure.

### Specificity analyses

To confirm whether the individual differences discovered in our analyses are specific to ADHD symptoms, we additionally tested other mental disorders, including depression, anxiety disorder, somatic problem, oppositional defiant disorder (ODD), and conduct disorder. These mental disorders, like ADHD, were assessed using CBCL. For each disorder, we tested the ATE estimate, model calibration, and performed the GATE test. These analyses help determine if individual difference patterns observed in ADHD symptoms are unique or if similar patterns exist across other mental health outcomes.

## Results

Our three-step model analysis culminated in a final optimized model that demonstrated an increase in ADHD symptoms following exposure to stressful events at the 1-year and 2-year follow-ups (1-year: ATE=1.29; 2-year: ATE=.94). Additionally, the final model revealed individual differences in these effects (1-year: Heterogeneity index=1.35; 2-year: Heterogeneity index=1.26) (Table 1). At baseline, the experience of stressful events was associated with increased ADHD symptoms (p < .05); however, the first model showed no significant variation among individuals (p > .05), eliminating the need for further testing (Table S3). The results of Random Iteration 1 are presented in Table 1 and the results of Random Iteration 2 and 3 are presented in Table S3.

**Table 1.** Final model (Random Iteration 1) results of average treatment effect and model calibration.

|  | ATE | FDR | Model fit | FDR | Heterogeneity index | FDR |
| --- | --- | --- | --- | --- | --- | --- |
| 1-year follow-up | 1.29 | 2.0E-09 | .99 | 1.6E-17 | 1.35 | .006 |
| 2-year follow-up | .94 | 5.5E-05 | .90 | 2.2E-08 | 1.26 | 2.0E-05 |

*Note.* A model fit and heterogeneity index close to 1 indicates a good fit. Detailed results of
Random Iteration 2 and 3 are presented in Table S3. ATE = average treatment effect.

At the 1-year follow-up, each stage of our three-step modeling process yielded significant results (p < .05; Table S3), resulting in a final model with three key covariates (i.e., parental depressive problems, parental ADHD, smoker PRS) (Figure 3.A) that showed consistent importance in the first and second step models (Figure S1). The GATE test revealed significant differences between the low and high-risk groups in the final model (low-risk(Q1): ATE=.87; high-risk(Q3): ATE=1.98; Q3-Q1: p FDR < .05) (Figure 3.B, Table S4), validating the individual differences. High-risk children exhibited higher levels of parental depressive problems and parental ADHD, but lower levels of polygenic scores for smoking compared to low-risk children (Figure 3.C, Figure S2). The partial dependence simulation confirmed this pattern, showing an increasing treatment effect for vulnerable factors (i.e., parental depressive problems and parental ADHD) and a decreasing effect for the resilient factor (i.e., smoking PRS) (Figure 3.D, Figure S2).

**Figure 3.**
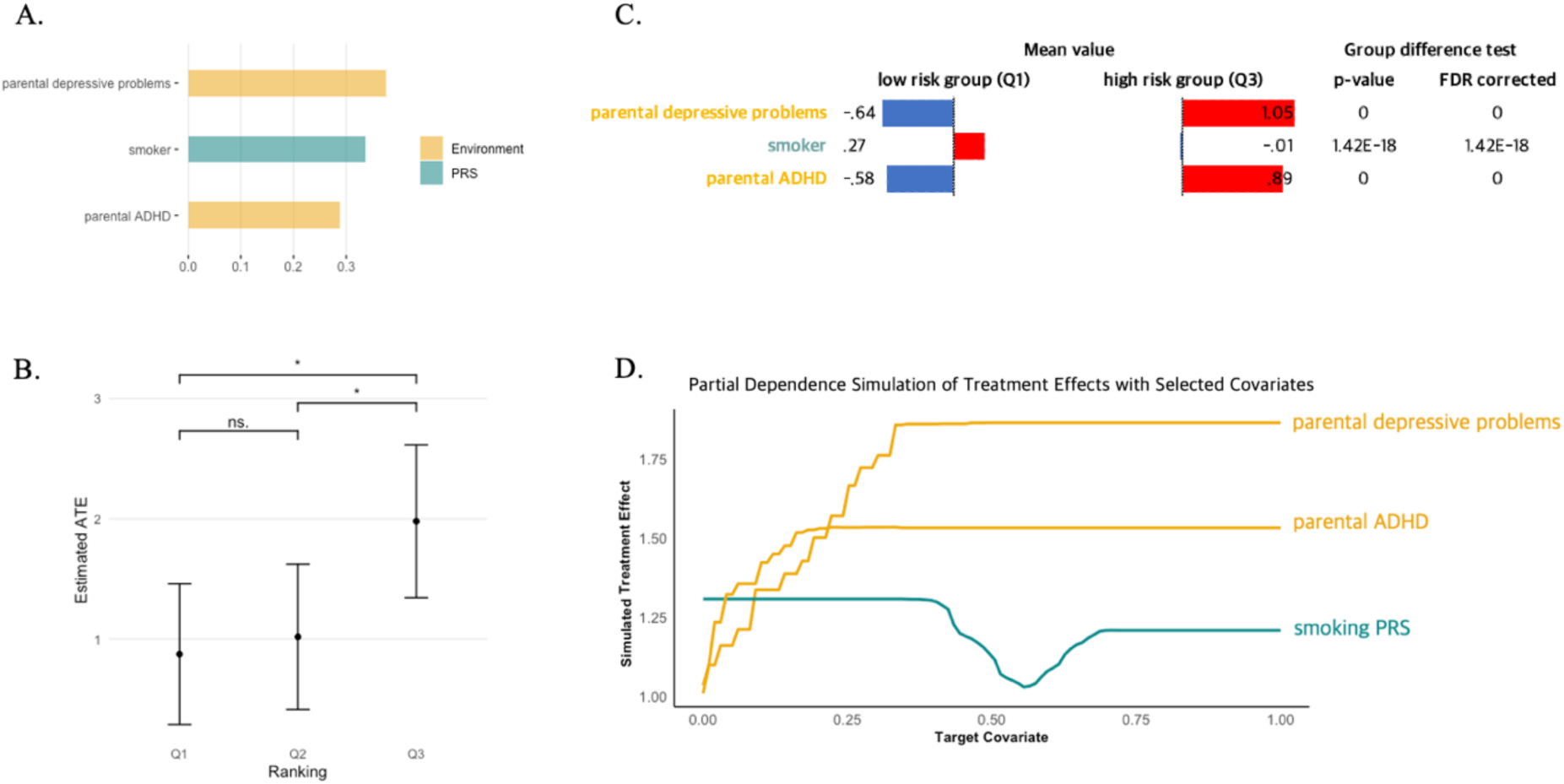
Final model (Random Iteration 1) results of the GATE test and covariate analyses at 1-year follow-up. A. Variable importance results. The x-axis refers to variable importance estimate. Yellow bar indicates environmental features and green for polygenic risk score. These three covariates were the variables that consistently showed above mean importance in the first and second models. B. The GATE test results. The results showed significant group differences between the high-risk and both the low- and middle-risk groups, suggesting the presence of individual differences on ADHD symptoms at 1-year follow-up following early-life stress (p FDR < .05). Q1 refers to low-risk group; Q2, middle-risk group; and Q3, high-risk group. C. The comparison results of covariate values between the low and high-risk groups. All covariate values are z-scaled. High-risk group showed more parental mental health problems but lower polygenic risk score for smoker than low-risk group. D. Partial dependence simulation results. The x-axis refers to the covariate value and y-axis refers to the simulated treatment effect of each covariate. The risk factors showed increasing treatment effect, whereas protective factor exhibited decreasing treatment effect when considering the multivariate interactions. Detailed results for all Random Iterations are presented in Figure S1, S3 and Table S4. ATE = average treatment effect; GATE = group average treatment effect; PRS = polygenic risk score; ns = not significant; *** p < .001; ** p < .01; * p < .05

At the 2-year follow-up, out of 55 initial covariates, the final model included six key covariates: parental internalizing problems, parental externalizing problems, parental depressive problems, parental antisocial personality problems, ADHD PRS, and streamline count between left precentral gyrus (PrCG) and right caudal anterior cingulate gyrus (CACG) (Figure 4.A, Figure S3). This final model demonstrated robustness through the GATE test, showing significant differences between the low and high-risk groups across all three Random Iterations (low risk(Q1): ATE=.08; high risk(Q2): ATE=1.99; Q3-Q1: p FDR < .01) (Figure 4.C, Table S5).

**Figure 4.**
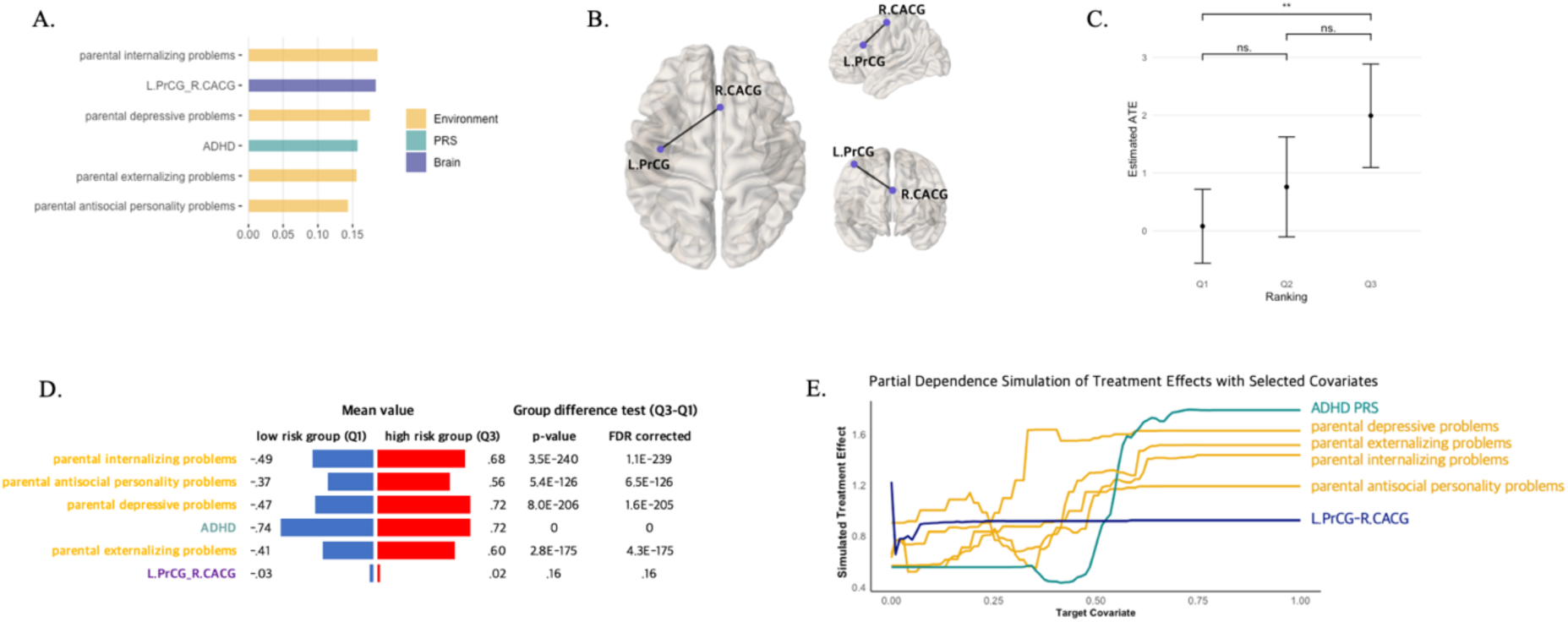
Final model (Random Iteration 1) results of the GATE test and covariate analyses at 2-year follow-up. A. Variable importance results. Yellow bar indicates environmental features; green, polygenic risk score, and blue for brain features. These six covariates were the variables that consistently showed above mean importance in the first and second models. B. The structural connectivity between left precentral gyrus (PrCG) and right caudal anterior cingulate gyrus (CACG). This connectivity was included as a covariate in the 2-year follow-up models. C. The GATE test results. The results showed significant group differences between the low- and high-risk groups, suggesting the presence of individual differences on ADHD symptoms at 2-year follow-up following early-life stress (p FDR < .05). Q1 refers to low-risk group; Q2, middle-risk group; and Q3, high-risk group. D. The comparison results of covariate values between the low and high-risk groups. All covariate values are z-scaled. High-risk group showed more parental mental health problems and higher polygenic risk score for ADHD than low-risk group. E. Partial dependence simulation results. The x-axis refers to the covariate value and y-axis refers to the simulated treatment effect of each covariate. Increase in simulated treatment effect was found for most risk factors, however, the streamline count between left PrCG and right CACG showed a slight initial increase but remained constant. Detailed outcomes for all Random Iterations are presented in Table S4, Figure S2, and S4. L.PrCG = left precentral gyrus; R.CACG = right caudal anterior cingulate gyrus; ATE = average treatment effect; GATE = group average treatment effect; PRS = polygenic risk score; ns = not significant; *** p < .001; ** p < .01; * p < .05

In assessing group differences in covariates, the high-risk group exhibited more parental mental health issues and a higher PRS for ADHD than the low-risk group (p FDR < .05) (Figure 4.D). However, the streamline count between the left PrCG and the right CACG varied across iterations (Figure S4). The partial dependence simulation showed a similar pattern. The treatment effect increased with higher ADHD PRS and parental mental illnesses (i.e., depressive, externalizing, internalizing, and antisocial personality problems) (Figure 4.E, Figure S4). However, the streamline count between the left PrCG and right CACG showed a slight initial increase but then remained constant, reporting the lowest treatment effect among the covariates. This finding may imply that the neural factor’s contribution to individual differences might not be as robust as other covariates, such as parental mental health problems and ADHD PRS.

### Specificity Analysis

To test whether our findings of individual differences in the impact of stressful events are specific to ADHD symptom, we applied the final model of 1-year and 2-year follow-ups to other mental disorder outcomes (i.e., depression, anxiety disorder, somatic problem, ODD, and conduct disorder). We conducted the GATE test for models showing both significant model fit and heterogeneity index; anxiety disorder and conduct disorder at 1-year follow-up, and depression, anxiety disorder, and ODD at 2-year follow-up (p < .05) (Table S6). As a result, these five models revealed no significant GATE results (Figure S5), underscoring the specificity of our model to ADHD symptom.

While some significant group differences were observed in the model with conduct disorder at 1-year follow-up, and with anxiety disorder and ODD at 2-year follow-up, the estimated ATE of the three groups did not show a consistent increase. This lack of monotonic increase indicates that the causal forest predictions did not align with the estimated ATEs, showing the specificity of our model to ADHD symptoms.

## Discussion

This study offers new insights into the multifaceted determinants of ADHD symptoms following stressful life events, highlighting the significance of parental influence, genetic predisposition, and neural factors in shaping children’s susceptibility or resilience. Notable individual differences emerged in ADHD symptom progression at 1-year and 2-year follow-ups, with such variations absent at baseline, suggesting the evolution of vulnerability or resilience factors over time. Our comprehensive assessment of environmental, genetic, and neural factors identified key contributors to these differences, underscoring the importance of personalized approaches in both research and clinical practice.

Initially, the impacts of stressful events on ADHD symptoms did not vary significantly among children; however, with time, marked individual differences became apparent. This trend indicates that children’s initial resilience to stress may not exert a strong influence on subsequent symptomatology, but as time progresses, these factors increasingly affect symptom trajectories, with certain children exhibiting heightened vulnerability. Literature supports this notion, demonstrating that the mental health outcomes for vulnerable children tend to deteriorate, while resilient children’s conditions typically stabilize or improve (Masten & Cicchetti, 2010; Masten & Tellegen, 2012).

The divergence in response to stress pertaining to ADHD symptoms is attributed to the interplay of neural, behavioral, and genetic factors, as our findings indicate. At the 1-year follow-up, children who were more susceptible to exhibiting increased ADHD symptoms upon exposure to stress were identified by the non-linear patterns among the several factors: higher instances of parental depressive problems and ADHD and a lower polygenic score of smoking. Surprisingly, a lower polygenic score for smoking emerged as a risk factor, a finding that defies conventional expectations.

Employing GRF method allowed us to uncover complex and multivariate relationships among variables that might be obscured when analyses focus narrowly on isolated factors. For example, a previous study discovered educational attainment and smoking initiation polygenic scores as primary predictors of psychiatric status in a clinical sample, utilizing regression analysis (Jansen et al., 2021). In contrast, our machine-learning strategy, adept at handling non-linear interactions among a wider array of variables, including parental depression and ADHD, has yielded novel insights. Notably, it surfaced the counterintuitive finding that lower polygenic scores for smoking could enhance vulnerability. Supporting this nuanced view, another study uncovered distinct patterns of susceptibility to post-traumatic stress symptoms after a natural disaster, including factors like higher education levels, lower family income, and significant depressive issues (Shiba et al., 2022). These findings collectively underscore the value of multivariate analytical approaches to fully understand the intricate dynamics of ADHD symptomatology following early-life stress.

By the 2-year follow-up, the vulnerability pattern shifted, encompassing a higher ADHD PRS, exacerbated parental mental health issues, and increased neural connectivity, particularly a greater white matter streamline count between the left PrCG and right CACG measured at baseline. This connection is part of the cognitive control network (CCN), which is crucial for attention, working memory, and executive functions (Cole & Schneider, 2007). Disrupted functional connectivity in the CCN has been reported in trauma-exposed veterans (Kennis, Rademaker, Van Rooij, Kahn, & Geuze, 2015) and ADHD adolescents (Francx et al., 2015). However, our results showed that the streamline count between the left PrCG and right CACG was not effective risk factor compared to other covariates. This may be due to the neurodevelopmental processes during adolescence, as higher-order cognitive functions like executive function continue to mature into adulthood (Best & Miller, 2010). Given that the mean age of participants in our study at baseline was 9.9 years, their structural connectivity between PrCG and CACG may still be maturing, making the differences between low and high-risk children less robust.

Although many children exhibit stress resilience by maintaining a capacity to buffer adversity’s impacts (Alvord & Grados, 2005; Zolkoski & Bullock, 2012), there are children who remain at heightened risk. Thus, the gene-brain-environmental patterns identified at the 2-year follow-up could be critical markers of sustained vulnerability reflecting an enduring susceptibility to ADHD symptoms over time. Unraveling how these intricate relationships evolve over time could lead to the development of more effective, personalized strategies to support children facing early-life stress and to prevent the exacerbation of ADHD symptoms.

The complex interactions between PRS (i.e., smoking, ADHD), brain structural connectivity, and environmental factors (i.e., parental mental illness) in the association between early-life stress and ADHD symptoms may be explained by epigenetic modifications highlighted in a prior study (Niwa et al., 2013). The study demonstrated that stress during critical developmental periods, such as adolescence, can induce lasting behavioral and neurobiological changes via glucocorticoid-mediated epigenetic control of dopaminergic neurons when combined with specific genetic risks (Niwa et al., 2013). Using a mouse model, they found that elevated glucocorticoids led to significant changes in dopaminergic projections from the ventral tegmental area only in the presence of both genetic predispositions and environmental stressors, highlighting a crucial interplay between genes and environment. These dopaminergic projections are involved in numerous brain functions, such as cognitive control and motivation (Cools, 2016), which is highly related to ADHD (Swanson et al., 2007). Thus, variability in ADHD symptom progression among children following stressful events can be partially explained by these findings. Indeed, a review on ADHD reported the associations with ADHD and dopamine genes, abnormal structure and function in brain regions related to dopamine (Swanson et al., 2007), along with significant effects of dopamine genes on brain anatomy (Durston, Mulder, Casey, Ziermans, & van Engeland, 2006). Due to the lack of physiological data in ABCD study, further study is necessary to disentangle the relationships between genes, brain, and environment from a molecular perspective, which could lead to more personalized approaches in managing ADHD symptoms after stress exposure.

This study marks a significant advancement in revealing the multifaceted relationships between stressful events and ADHD symptoms in children. Utilizing GRF method, a flexible non-parametric algorithm, we illustrate how early-life stress not only exacerbates ADHD symptoms but also leads to diverging symptom trajectories over time. Moreover, we identified key factors—ranging from genetic predispositions to brain connectivity—that influence whether a child might become more vulnerable to or resilient against developing ADHD after experiencing stress. To our knowledge, this is the first study to show such variability in a longitudinal design, revealing the intricate interaction between genetic, neural, and environmental factors. Still, there are limitations to consider. Our operational definition of stressful life events, based on exposure, may not adequately capture the subjective intensity of these experiences, pointing to a direction for future research. The lack of data in ABCD dataset precluded the inclusion of subjective perceptions, highlighting an avenue for future research to explore individual variances not captured in our study. Additionally, GRF results might vary with changes in seed numbers or the ordering of covariates. We addressed this by performing multiple iterations with different seed numbers and sequences of covariates, complemented by a three-step model analysis to ensure robustness (i.e., first model, second model, final model). Lastly, the study’s scope, while comprehensive, does not extend into the potential long-term impacts of early-life stress into adolescence and adulthood, which warrants further investigation.

## Key points

- This study investigates how early-life stress affects children’s development of ADHD symptoms, emphasizing individual variability.
- Using a novel non-parametric machine-learning approach, we identified key factors, such as parental mental health issues, ADHD PRS, and brain connectivity, that influence whether a child might become more vulnerable to or resilient against developing ADHD after experiencing stress.
- Identifying key risk factors can help clinicians tailor interventions to children at higher risk for developing ADHD following stress.
- This research underscores the importance of considering multifaceted gene-brain-environment interactions in future studies of ADHD and stress, advocating for more sophisticated analytical approaches to uncover these complex dynamics.

The authors declare no conflicts of interest.

## Data availability statement

Data were obtained from ABCD study (http://abcdstudy.org). To access, users are required to register for an account via the NIMH Data Archive and adhere to the instructions provided on the website to secure permission. GRF algorithm code can be found at https://grf-labs.github.io/grf/index.html.

## Acknowledgements

This work was supported by the National Research Foundation of Korea(NRF) grant funded by the Korea government(MSIT) (No. 2021R1C1C1006503, RS-2023-00266787, RS-2023-00265406, RS-2024-00421268), by Creative-Pioneering Researchers Program through Seoul National University(No. 200-20230058), by Semi-Supervised Learning Research Grant by SAMSUNG(No.A0426-20220118), by Identify the network of brain preparation steps for concentration Research Grant by LooxidLabs(No.339-20230001), by Institute of Information & communications Technology Planning & Evaluation (IITP) grant funded by the Korea government(MSIT) [NO.RS-2021-II211343, Artificial Intelligence Graduate School Program (Seoul National University)] and by the National Supercomputing Center with supercomputing resources including technical support(KSC-2023-CRE-0568) and by the Ministry of Education of the Republic of Korea and the National Research Foundation of Korea (NRF-2021S1A3A2A02090597).

## Notes

### Competing Interest Statement

The authors have declared no competing interest.

### Author Declarations

Data in this study were obtained from the Adolescent Brain Cognitive Development (ABCD) Study 4.0 release (https://abcdstudy.org/). The ABCD Study manages participant consent and assent procedures, with its protocols receiving approval from a centralized Institutional Review Board at the University of California, San Diego.

