## Supplementary Information for "Individual Differences in Effects of Stressful Life Events on Childhood ADHD: Genetic, Neural, and Familial Contributions"

### Supporting Information

#### Appendix S1.

Covariate measurements. ……………………………………………………………………………2

#### Supplementary Results

**Table S1.** Genome-wide association studies used for polygenic risk score generation………………6

**Table S2.** The results of regression analyses to extract ADHD-specific brain features………….…. 7

**Table S3.** The three-step model results in the impact of stressful events on ADHD symptoms at baseline, 1-year follow-up, and 2-year follow-up ….…………………………………………..…8

**Figure S1.** Variable importance of three-step models in the impact of stressful events on ADHD symptoms at 1-year follow-up...……………………………...…………………………………....9

**Table S4.** The GATE test results of final model evaluating the impact of stressful events on ADHD symptoms at 1-year follow-up………………………………………………………………....13

**Figure S2.** The results of group difference test and partial dependence simulation at 1-year follow-up ………………………………………………………………………...…………………14

**Figure S3.** Variable importance of three-step models in the impact of stressful events on ADHD symptoms at 2-year follow-up ..……………………………...……………………………………..11

**Table S5.** The GATE test results of final model evaluating the impact of stressful events on ADHD symptoms at 2-year follow-up………………………………………………………………....13

**Figure S4.** The results of group difference test and partial dependence simulation at 2-year follow-up ……………………………………………………………………………………………...15

**Table S6.** The final model results of specificity analyses in the impact of stressful events on different mental disorders at 1-year and 2-year follow-ups ……...…………...…………………... 16

**Figure S5.** The GATE test of specificity analyses in the impact of stressful events on different mental disorders at 1-year and 2-year follow-ups………...……………………………………… 17

#### Appendix S1. Covariate measurements

**Sociodemographic variables.** The sociodemographic factors consist of the child's characteristics and environment of family, neighborhood, and school. Child characteristics encompass age, sex, ethnicity (i.e., White, Black, Hispanic, Asian, Other), BMI, birth country (i.e., US or not US), abuse, neglect, and intelligence quotient (IQ). Abuse assessment utilized the parent-reported KSADS-PTSD, covering emotional, sexual, and physical abuse dimensions. Following a prior study (Stinson et al., 2021), neglect is categorized into physical and emotional, with the former assessed via the Parental Monitoring scale (Chilcoat & Anthony, 1996) and the latter through the Child Report of Parent Behaviors Inventory (Schaefer, 1965). IQ measurements were obtained using the NIH Toolbox Cognition total composite score (Akshoomoff et al., 2014).

Family environment included parental age, marital status (i.e., married, widowed, divorced, separated, never married, living with partner), parental education level (i.e., highest grade or level of school you have completed or highest degree you have received), family income (i.e., total combined family income for past 12 months), family environment, family financial adversity, family depression history, and parental mental health. The family environment was assessed using the youth-reported Family Environment Scale-Family Conflict Subscale Modified from PhenX (Moos & Moos, 1994), where higher scores indicate increased family conflict. The Parent-Reported Financial Adversity Questionnaire (PRFQ) (Diemer, Mistry, Wadsworth, López, & Reimers, 2013) assessed financial adversity, querying the family's capability to afford basic living expenses, with higher scores denoting severe financial hardships. Family depression history was determined by caregiver reports, using a 4-level assessment based on depressive histories across two generations (van Dijk, Murphy, Posner, Talati, & Weissman, 2021), categorizing risk from lowest (neither generation affected) to highest (both generations affected). Parental mental health was measured using the Adult Self-report (ASR) (Achenbach & Rescorla, 2003), which includes a range of mental health problems with higher scores indicating greater issues (e.g., externalizing behavior problems, internalizing behavior problems, depressive problems, anxiety problems, somatic problems, avoidant personality problems, ADHD, and antisocial personality problems).

Neighborhood environment included rates of total violent and drug offenses in the child's neighborhood, assessed via the Uniform Crime Report from the Federal Bureau of Investigation, distributed by Inter-university Consortium for Political and Social Research (United States Department Of Justice. Office Of Justice Programs. Federal Bureau Of Investigation, 2012). The school environment was measured using the school environment subscale from the School Risk and Protective Factors questionnaire (Arthur et al., 2007), which evaluates the child's perception of school climate, with higher scores reflecting a more positive environment.

**Genetic features.** To create the polygenic risk scores (PRSs), saliva samples were collected from study participants and genotyped using Affymetrix SmokeScreen array, which looks at 733,293 single nucleotide variations (SNVs). To ensure the quality of genotype data, certain SNVs that had low genotype call rate (< 95%), low sample call rate (< 95%), and low minor allele frequency (< 1%) were excluded. Genotypes were then imputed from the 1000 Genomes Project phase3 reference panel (Consortium, 2015) using the Michigan Imputation Server (Conomos, Miller, & Thornton, 2015) and phased using the Eagle version 2.4 (Loh et al., 2016), which included a total of 12,046,090 SNVs. Further quality control procedures were applied to remove SNVs with low imputation quality scores (INFO score <0.4), low genotype call rate (< 95%), high sample missingness (> 5%), low minor allele frequency (< 0.5%), a Hardy-Weinberg equilibrium p-value below $1 \times 10^{-20}$, and extreme heterozygosity (population mean>±3SD). These procedures remained 11,221,810 variants.

To account for diverse ethnic backgrounds and genetic ancestries among ABCD study participants, we implemented a rigorous QC process to mitigate potential population stratification from genetic relatedness and ancestry admixture. We used the SNPRelate R package (Zheng et al., 2012) to identify genetically unrelated individuals and computed ancestrally informative principal components (PCs). Employing the PC-Air algorithm, two rounds of principal component analysis (PCA) robust against familial or cryptic relatedness were conducted. Initially, pairwise kinship coefficients were estimated using the KING-robust algorithm with a pruned set of independent genetic variants (LD threshold r-squared < 0.1). We excluded individuals related closer than third-degree relatives, designating the excluded as a held-out validation set (n=1,814). PC-Air then processed 8,845 unrelated individuals, recalculating kinship using PC-Relate adjusted for ancestry. A second

PCA refined the PCs for these individuals. We removed 88 participants whose projected PCs showed Mahalanobis distances greater than 6SDs. Ultimately, among 8,620 unrelated individuals we used 6,555 European ancestry individuals as the final baseline set for main analysis.

The genetic features include PRS of 17 traits that can be classified as three domains (i.e., cognitive traits, mental disorders, substance use): educational attainment (EA) (Lee et al., 2018), cognitive performance (CP) (Lee et al., 2018), IQ (Savage et al., 2018), depression (Howard et al., 2019), anxiety (Otowa et al., 2016), neuroticism (Nagel et al., 2018), worry (Nagel et al., 2018), ADHD (Demontis et al., 2019), autism spectrum disorder (ASD) (Grove et al., 2019), bipolar disorder (Stahl et al., 2019), major depressive disorder (MDD) (Wray et al., 2018), PTSD (Bierut, Nelson, Rice, Saccone, & Heath, 2019), schizophrenia (Ruderfer et al., 2018), subjective well-being (Okbay et al., 2016), general happiness (http://www.nealelab.is/ukbiobank/), alcoholic drinks per week (Karlsson Linnér et al., 2019), and smoking status (Karlsson Linnér et al., 2019). We used publicly available summary statistics of genome-wide association studies (GWAS), and the details of GWAS summary statistics collected for polygenic scoring are listed in Table S1. For polygenic scoring method, PRS-CS (Ge, Chen, Ni, Feng, & Smoller, 2019), a Bayesian polygenic modeling technique, was employed. The method is known for its effective posterior inference algorithm considering allele frequencies and linkage disequilibrium (LD) patterns by adopting continuous shrinkage prior. The LD reference panel of European from the 1000 Genome Project phase 3 was utilized in accordance with the discovery GWAS sample for PRS construction. Though PRS-CS could automatically estimate the parameter, we applied small-scale grid search of global shrinkage parameter as recommended in the original paper for whose related phenotype is available within the ABCD study for better predictive performance. In this way, we optimized PRSs of 8 traits (depression, MDD, ADHD, Subjective well-being, PTSD, CP, EA, IQ) by choosing the optimal global shrinkage hyperparameter in an independent held-out validation set of 1,579 unrelated participants, which were removed during the QC process of relatedness analysis. For the 9 GWAS traits that do not have target outcome variables available in the ABCD study, we performed pseudo-validation using PRS-CS-auto, in which the hyperparameter is automatically selected from data with a fully Bayesian approach. All final GPSs were residualized by top 10 genetic PCs.

**Brain features.** Following process were applied to preprocess diffusion MRI data (Kim et al., 2022): eddy current distortion correction (Zhuang et al., 2006), head motion correction (Hagler et al., 2009), diffusion gradient adjustment for head rotation (Hagler et al., 2009; Leemans & Jones, 2009), diffusion tensor model fitting (Chang, Jones, & Pierpaoli, 2005), b0 distortion correction (Holland, Kuperman, & Dale, 2010), and gradient nonlinearity distortion correction (Jovicich et al., 2006). The data were obtained using mutual information of T2-weighted b0 images to T1w structural images (Wells III, Viola, Atsumi, Nakajima, & Kikinis, 1996) and then resampled to a standard orientation with 1.77 mm isotropic resolution.

We used individual connectome data to obtain precise brain imaging measurements. We first conducted whole-brain white matter tracts assessment and individualized connectome creation using MRtrix3 software (Tournier et al., 2019). For connectivity analysis, streamline counts associated with both fiber connection strength (Cha et al., 2015, 2016) and fiber integrity were employed. We used spatially varying noise maps to determine the threshold for PCA denoising based on the noise level (Veraart et al., 2016), and bias correction was performed using the N4 algorithm in the Advanced Normalization Tools (ANTs) pipeline (Tustison et al., 2010). We obtained a connectivity index with a white matter pathway (Ciccarelli et al., 2006) by performing a probabilistic tractography. Using a final streamline count of 10 million, we created an 84x84 whole-brain connectome matrix for each participant based on T1-based parcellation and segmentation from FreeSurfer.

**Table S1. Genome-wide association studies used for polygenic risk score generation.**

| Trait | GWAS sample size | Study |
|---|---|---|
| Educational attainment | 1,131,881 | Lee JJ, Wedow R, Okbay A, et al. Gene discovery and polygenic prediction from a genome-wide association study of educational attainment in 1.1 million individuals. *Nature Genetics*. 2018-08-01 2018;50(8):1112-1121. doi:10.1038/s41588-018-0147-3 |
| Cognitive performance | 1,131,881 | Lee JJ, Wedow R, Okbay A, et al. Gene discovery and polygenic prediction from a genome-wide association study of educational attainment in 1.1 million individuals. *Nature Genetics*. 2018-08-01 2018;50(8):1112-1121. doi:10.1038/s41588-018-0147-3 |
| IQ | 269,867 | Savage JE, Jansen PR, Stringer S, et al. Genome-wide association meta-analysis in 269,867 individuals identifies new genetic and functional links to intelligence. *Nature Genetics*. 2018-07-01 2018;50(7):912-919. doi:10.1038/s41588-018-0152-6 |
| Depression | 500,199 | Howard DM, Adams MJ, Clarke T-K, et al. Genome-wide meta-analysis of depression identifies 102 independent variants and highlights the importance of the prefrontal brain regions. *Nature Neuroscience*. 2019-03-01 2019;22(3):343-352. doi:10.1038/s41593-018-0326-7 |
| Anxiety | 31,060 | Otowa, T., Hek, K., Lee, M., Byrne, E. M., Mirza, S. S., Nivard, M. G., ... & Hettema, J. M. (2016). Meta-analysis of genome-wide association studies of anxiety disorders. *Molecular psychiatry*, *21*(10), 1391-1399. |
| Neuroticism | 390,278 | Nagel M, Jansen PR, Stringer S, et al. Meta-analysis of genome-wide association studies for neuroticism in 449,484 individuals identifies novel genetic loci and pathways. *Nature Genetics*. 2018-07-01 2018;50(7):920-927. doi:10.1038/s41588-018-0151-7 |
| Worry | 348,219 | Nagel M, Jansen PR, Stringer S, et al. Meta-analysis of genome-wide association studies for neuroticism in 449,484 individuals identifies novel genetic loci and pathways. *Nature Genetics*. 2018-07-01 2018;50(7):920-927. doi:10.1038/s41588-018-0151-7 |
| ADHD | 53,293 | Demontis D, Walters RK, Martin J, et al. Discovery of the first genome-wide significant risk loci for attention deficit/hyperactivity disorder. *Nature Genetics*. 2019-01-01 2019;51(1):63-75. doi:10.1038/s41588-018-0269-7 |
| ASD | 46,350 | Grove J, Ripke S, Als TD, et al. Identification of common genetic risk variants for autism spectrum disorder. *Nature Genetics*. 2019-03-01 2019;51(3):431-444. doi:10.1038/s41588-019-0344-8 |
| Bipolar disorder | 51,710 | Stahl EA, Breen G, Forstner AJ, et al. Genome-wide association study identifies 30 loci associated with bipolar disorder. *Nature Genetics*. 2019-05-01 2019;51(5):793-803. doi:10.1038/s41588-019-0397-8 |
| MDD | 480,359 | Wray NR, Ripke S, Mattheisen M, et al. Genome-wide association analyses identify 44 risk variants and refine the genetic architecture of major depression. *Nature Genetics*. 2018-05-01 2018;50(5):668-681. doi:10.1038/s41588-018-0090-3 |
| PTSD | 174,659 | Nievergelt CM, Maihofer AX, Klengel T, et al. International meta-analysis of PTSD genome-wide association studies identifies sex- and ancestry-specific genetic risk loci. *Nature Communications*. 2019-12-01 2019;10(1)doi:10.1038/s41467-019-12576-w |
| Schizophrenia | 65,967 | Ruderfer DM, Ripke S, Mcquillin A, et al. Genomic Dissection of Bipolar Disorder and Schizophrenia, Including 28 Subphenotypes. *Cell*. 2018-06-01 2018;173(7):1705-1715.e16. doi:10.1016/j.cell.2018.05.046 |
| Subjective well-being | 128,049 | Okbay A, Baselmans BML, De Neve J-E, et al. Genetic variants associated with subjective well-being, depressive symptoms, and neuroticism identified through genome-wide analyses. *Nature Genetics*. 2016-06-01 2016;48(6):624-633. doi:10.1038/ng.3552 |
| General happiness | 125,527 | UK Biobank GWAS. Neale Lab. http://www.nealelab.is/ukbiobank/ Accessed Apr 29. 2020. |
| Drinking | 46,568 | Karlsson Linnér R, Biroli P, Kong E, et al. Genome-wide association analyses of risk tolerance and risky behaviors in over 1 million individuals identify hundreds of loci and shared genetic influences. *Nature Genetics*. 2019-02-01 2019;51(2):245-257. doi:10.1038/s41588-018-0309-3 |
| Smoking status | 518,633 | Karlsson Linnér R, Biroli P, Kong E, et al. Genome-wide association analyses of risk tolerance and risky behaviors in over 1 million individuals identify hundreds of loci and shared genetic influences. *Nature Genetics*. 2019-02-01 2019;51(2):245-257. doi:10.1038/s41588-018-0309-3 |

All traits are European ancestry. ASD = autism spectrum disorder; MDD = major depressive disorder; PTSD = post-traumatic stress disorder

**Table S2. The results of regression analyses to extract ADHD-specific brain features.**

| ROI | Estimate | p-value | FDR |
|---|---|---|---|
| **Baseline** | | | |
| left caudal anterior cingulate (surface area) | -.00141 | 3.72E-03 | .039 |
| left cuneus (surface area) | -.00114 | 1.06E-03 | .016 |
| left lingual (surface area) | -.00087 | 2.02E-06 | .001 |
| left pars orbitalis (surface area) | -.00281 | 3.50E-04 | .010 |
| left pars triangularis (surface area) | -.00130 | 5.37E-05 | .005 |
| left pericalcarine (surface area) | -.00100 | 9.17E-04 | .016 |
| left posterior cingulate (surface area) | -.00113 | 3.20E-03 | .035 |
| left rostral middle frontal (surface area) | -.00039 | 8.57E-05 | .006 |
| right caudal anterior cingulate (surface area) | -.00175 | 1.62E-04 | .008 |
| right cuneus (surface area) | -.00105 | 2.23E-03 | .026 |
| right lateral orbital frontal (surface area) | -.00075 | 1.09E-03 | .016 |
| right lingual (surface area) | -.00055 | 1.30E-03 | .018 |
| right pars orbitalis (surface area) | -.00248 | 1.29E-04 | .007 |
| right pericalcarine (surface area) | -.00098 | 7.16E-04 | .014 |
| right rostral anterior cingulate (surface area) | -.00226 | 2.22E-04 | .009 |
| right rostral middle frontal (surface area) | -.00028 | 1.52E-03 | .020 |
| right superior temporal (surface area) | -.00057 | 7.17E-04 | .014 |
| left lingual (volume) | -.00029 | 1.38E-05 | .002 |
| left pars orbitalis (volume) | -.00065 | 7.31E-04 | .014 |
| left pars triangularis (volume) | -.00031 | 1.13E-03 | .016 |
| left rostral middle frontal (volume) | -.00011 | 2.47E-04 | .009 |
| right caudal anterior cingulate (volume) | -.00044 | 6.08E-04 | .014 |
| right lateral orbital frontal (volume) | -.00027 | 2.80E-04 | .009 |
| right lingual (volume) | -.00021 | 6.41E-04 | .014 |
| right pars orbitalis (volume) | -.00053 | 9.66E-04 | .016 |
| right pericalcarine (volume) | -.00046 | 2.12E-03 | .025 |
| right rostral anterior cingulate (volume) | -.00048 | 2.06E-03 | .025 |
| left frontal pole – right transverse temporal (count) | 22.61 | 2.76E-06 | .010 |
| left frontal pole – right transverse temporal (FA) | 42.08 | 6.36E-06 | .022 |
| **1-year follow-up** | | | |
| left superior parietal – right posterior cingulate (count) | .016 | 7.60E-06 | .013 |
| left frontal pole – right transverse temporal (count) | 23.21 | 1.46E-06 | .005 |
| left frontal pole – right transverse temporal (FA) | 43.40 | 2.91E-06 | .010 |
| **2-year follow-up** | | | |
| left precentral – right caudal anterior cingulate (count) | .051 | 1.13E-08 | 4.0E-05 |
| right lateral occipital – right posterior cingulate (count) | .069 | 2.70E-08 | 4.7E-05 |
| right pars opercularis – right precuneus (count) | -.060 | 1.43E-05 | .017 |

Note. ROI = region of interest.

**Table S3. The three-step model results in the impact of stressful events on ADHD symptoms at baseline, 1-year follow-up, and 2-year follow-up**

| Model | ATE | p-value | FDR | Model fit | p-value | FDR | Heterogeneity index | p-value | FDR |
|---|---|---|---|---|---|---|---|---|---|
| **Baseline** | | | | | | | | | |
| First model (71 covariates) | | | | | | | | | |
| Random iteration 1 | 1.19 | 3.37E-10 | - | 1.02 | 6.35E-12 | - | .45 | .17 | - |
| Random iteration 2 | 1.04 | 1.55E-08 | - | .96 | 1.46E-11 | - | -.09 | .56 | - |
| Random iteration 3 | 1.06 | 5.28E-09 | - | .98 | 1.37E-10 | - | .52 | .13 | - |
| **1-year follow-up** | | | | | | | | | |
| First model (55 covariates) | | | | | | | | | |
| Random iteration 1 | 1.12 | 1.32E-08 | 1.98E-08 | 1.03 | 2.15E-12 | 3.37E-12 | .89 | .03 | .026 |
| Random iteration 2 | 1.12 | 1.60E-08 | 1.98E-08 | 1.05 | 2.66E-12 | 3.37E-12 | .98 | .01 | .024 |
| Random iteration 3 | 1.10 | 1.98E-08 | 1.98E-08 | 1.03 | 3.37E-12 | 3.37E-12 | 1.02 | .02 | .024 |
| Second model (12 covariates) | | | | | | | | | |
| Random iteration 1 | 1.04 | 8.26E-08 | 8.26E-08 | .98 | 3.25E-11 | 4.88E-11 | .95 | 4.26E-04 | .001 |
| Random iteration 2 | 1.11 | 1.06E-08 | 3.17E-08 | .97 | 2.41E-11 | 4.88E-11 | 1.36 | 8.55E-04 | .001 |
| Random iteration 3 | 1.07 | 4.12E-08 | 6.18E-08 | .92 | 3.08E-10 | 3.08E-10 | 1.78 | 2.54E-03 | .003 |
| Final model (3 covariates) | | | | | | | | | |
| Random iteration 1 | 1.29 | 2.00E-09 | 2.00E-09 | .99 | 1.56E-17 | 1.56E-17 | 1.35 | 4.93E-03 | .006 |
| Random iteration 2 | 1.29 | 1.01E-09 | 1.52E-09 | 1.00 | 2.64E-18 | 7.01E-18 | 1.12 | 5.50E-03 | .006 |
| Random iteration 3 | 1.31 | 5.10E-10 | 1.52E-09 | 1.00 | 4.67E-18 | 7.01E-18 | 1.17 | 3.62E-03 | .006 |
| **2-year follow-up** | | | | | | | | | |
| First model (55 covariates) | | | | | | | | | |
| Random iteration 1 | 1.01 | 8.32E-06 | 9.42E-06 | 1.02 | 2.74E-08 | 4.12E-08 | 1.12 | .03 | .03 |
| Random iteration 2 | 1.01 | 6.56E-06 | 9.42E-06 | 1.01 | 5.21E-08 | 5.21E-08 | .77 | .01 | .03 |
| Random iteration 3 | 1.00 | 9.42E-06 | 9.42E-06 | 1.00 | 2.10E-08 | 4.12E-08 | 1.10 | .03 | .03 |
| Second model (16 covariates) | | | | | | | | | |
| Random iteration 1 | 1.01 | 9.51E-06 | 1.24E-05 | .99 | 3.87E-09 | 5.81E-09 | 1.02 | 4.93E-03 | .005 |
| Random iteration 2 | 1.00 | 1.24E-05 | 1.24E-05 | 1.03 | 3.74E-09 | 5.81E-09 | .91 | 4.53E-04 | .001 |
| Random iteration 3 | 1.02 | 9.57E-06 | 1.24E-05 | 1.03 | 2.13E-08 | 2.13E-08 | 1.06 | 2.83E-04 | .001 |
| Final model (6 covariates) | | | | | | | | | |
| Random iteration 1 | .94 | 5.16E-05 | 5.48E-05 | .90 | 2.16E-08 | 2.16E-08 | 1.26 | 1.95E-05 | 1.95E-05 |
| Random iteration 2 | .94 | 5.48E-05 | 5.48E-05 | .90 | 1.13E-08 | 1.70E-08 | 1.35 | 1.72E-05 | 1.95E-05 |
| Random iteration 3 | 1.03 | 1.12E-05 | 3.37E-05 | .86 | 3.70E-09 | 1.11E-08 | 1.56 | 9.69E-06 | 1.95E-05 |

Note. A model fit and heterogeneity index close to 1 indicates a good fit. ATE = average treatment effect.

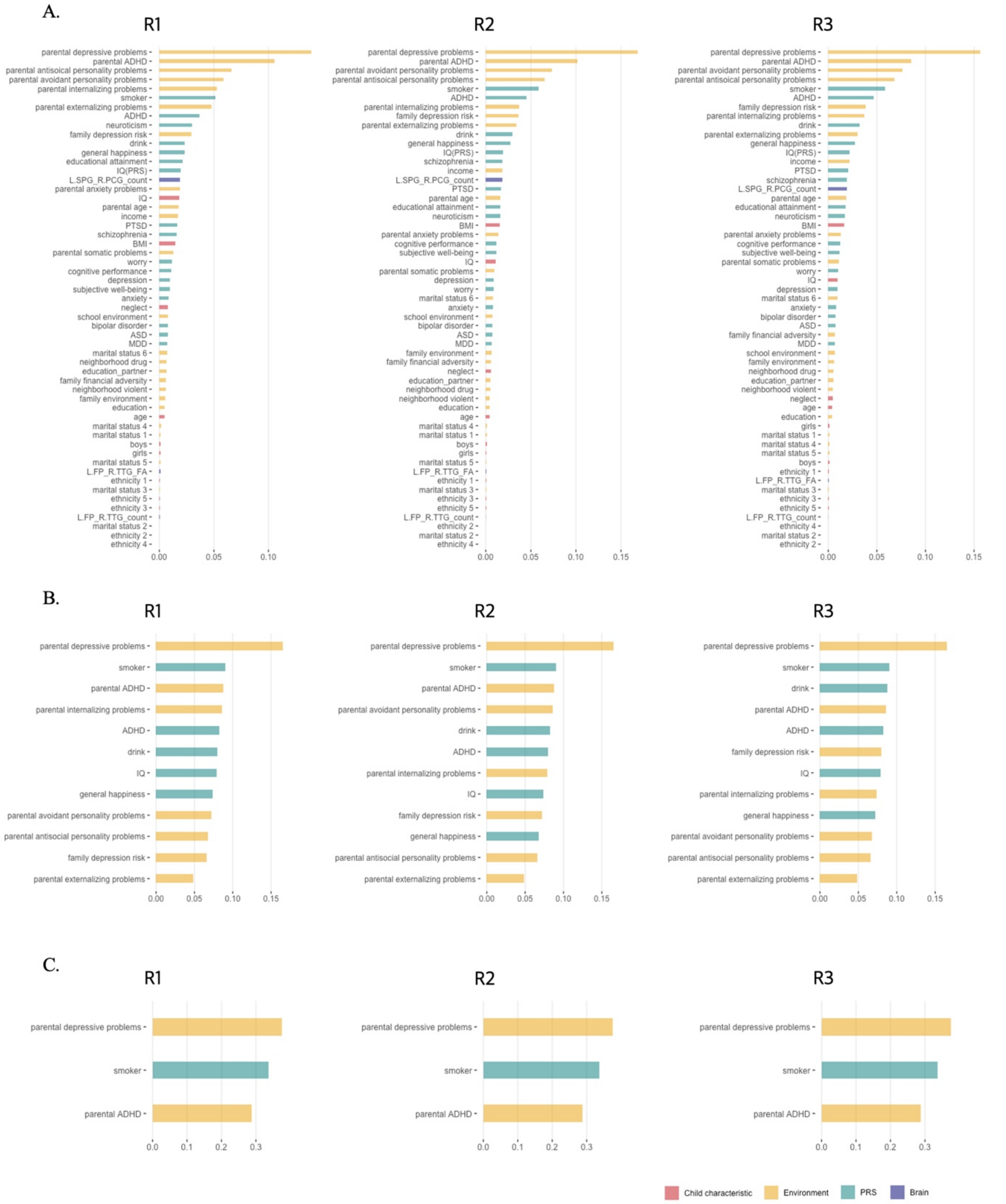


**Figure S1. Variable importance of three-step models in the impact of stressful events on ADHD symptoms at 1-year follow-up**

A. Variable importance of first model. This model incorporated 55 covariates. Variables demonstrating consistently higher-than-average importance across all three Random Iterations were identified and selected to form the 'second covariates' set for the second model.

B. Variable importance of second model. This stage involved analyzing the 12 covariates previously identified for their above-average importance from the first model. Among these, parental depressive problems, parental ADHD, and smoker PRS were consistently ranked with higher-than-average importance across the three Random Iterations. These variables were thus chosen as the 'final covariates' set for the final model.

C. Variable importance of final model. In the final model, the analysis focused on the three covariates identified in the second model.

Red bar indicates child characteristics; yellow bar, environmental factors; green bar, polygenic risk score; and blue bar, brain features.

R1 = random iteration 1; R2 = random iteration 2; R3 = random iteration 3; PRS = polygenic risk score; PTSD = post-traumatic stress disorder; BMI = body mass index; MDD = major depressive disorder; ASD = autism spectrum disorder; Marital status 1 = married; Marital status 2 = widowed; Marital status 3 = divorced; Marital status 4 = separated; Marital status 5 = never married; Marital status 6 = living with partner; Ethnicity 1 = White; Ethnicity 2 = Black; Ethnicity 3 = Hispanic; Ethnicity 4 = Asian; Ethnicity 5 = Other; L.SPG = left superior parietal gyrus; R.PCG = right posterior cingulate gyrus; L.FP = left frontal pole; R.TTG = right transverse temporal

**Table S4. The GATE test results of final model evaluating the impact of stressful events on ADHD symptoms at 1-year follow-up**

| | GATE estimate (SE) of each quantile | | | Group difference test (Q3-Q1) | |
|---|---|---|---|---|---|
| | Q1 | Q2 | Q3 | p-value | FDR |
| Random Iteration 1 | .87 (.29) | 1.02 (.30) | 1.98 (.32) | .01 | .03 |
| Random Iteration 2 | .84 (.24) | 1.18 (.36) | 1.16 (.31) | .01 | .04 |
| Random Iteration 3 | .84 (.26) | 1.86 (.34) | 1.91 (.29) | .006 | .02 |

Note. GATE = group average treatment effect; SE = standard error; Q1 = low-risk group; Q2 = middle-risk group; Q3 = high-risk group.

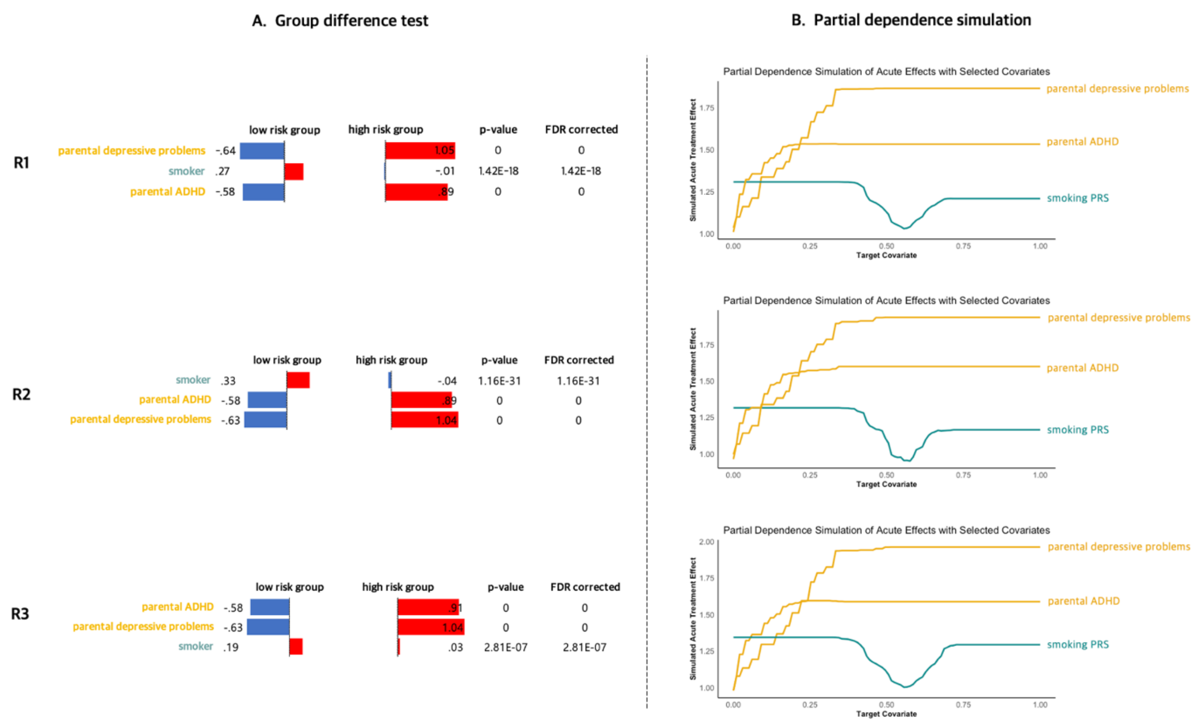


**Figure S2. The results of group difference test and partial dependence simulation at 1-year follow-up**

A. The comparison of three covariate values between low and high-risk groups across three Random Iterations. All covariate values are z-scaled. The high-risk group significantly exhibited more parental depressive problems and ADHD and lower polygenic scores of smoking for developing ADHD.

B. The results of partial dependence simulation across three Random Iterations. The x-axis refers to the covariate value and y-axis refers to the simulated treatment effect of each covariate. The risk factors showed increasing treatment effect, whereas protective factor exhibited decreasing treatment effect when considering the multivariate interactions.

Yellow indicates environmental factors and green for polygenic risk score.

R1 = random iteration 1; R2 = random iteration 2; R3 = random iteration 3; PRS = polygenic risk score.

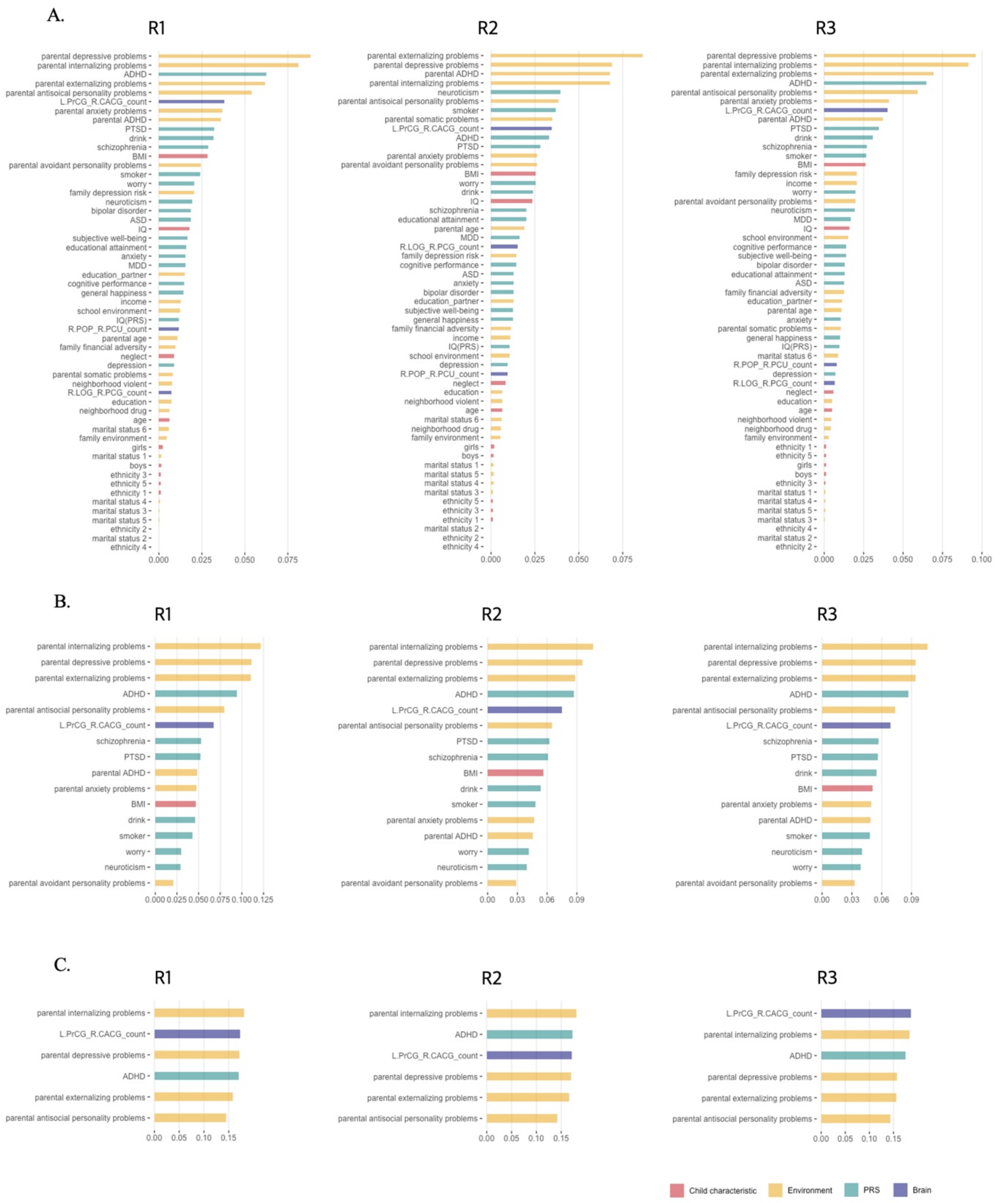


**Figure S3. Variable importance of three-step models in the impact of stressful events on ADHD symptoms at 2-year follow-up**

A. Variable importance of first model. This phase analyzed 55 covariates. The variables consistently showing higher-than-average importance across all three Random Iterations were identified and compiled into the 'second covariates' set for the second model.

B. Variable importance of second model. This stage involved 16 covariates that demonstrated above-average importance in the first model. The variables related to parental mental illnesses, ADHD PRS, and a neural factor were distinguished for their significance. These were advanced as the 'final covariates' set for the final model.

C. Variable importance of final model. In the final model, the analysis focused on the six covariates identified in the second model.

Red bar indicates child characteristics; yellow, environmental factors; green, polygenic risk score; and blue, brain features.

R1 = random iteration 1; R2 = random iteration 2; R3 = random iteration 3; PRS = polygenic risk score; BMI = body mass index; PTSD = post-traumatic stress disorder; MDD = major depressive disorder; ASD = autism spectrum disorder; Marital status 1 = married; Marital status 2 = widowed; Marital status 3 = divorced; Marital status 4 = separated; Marital status 5 = never married; Marital status 6 = living with partner; Ethnicity 1 = White; Ethnicity 2 = Black; Ethnicity 3 = Hispanic; Ethnicity 4 = Asian; Ethnicity 5 = Other; L.PrCG = left precentral gyrus; R.CACG = right caudal anterior cingulate gyrus.

**Table S5. The GATE test results of final model evaluating the impact of stressful events on ADHD symptoms at 2-year follow-up**

| | GATE estimate (SE) of each quantile | | | Group difference test (Q3-Q1) | |
|---|---|---|---|---|---|
| | Q1 | Q2 | Q3 | p-value | FDR |
| Random Iteration 1 | .08 (.32) | .76 (.43) | 1.99 (.45) | .0005 | .002 |
| Random Iteration 2 | .33 (.29) | .71 (.32) | 1.78 (.38) | .003 | .007 |
| Random Iteration 3 | .28 (.31) | .63 (.39) | 2.18 (.45) | .0005 | .001 |

Note. GATE = group average treatment effect; SE = standard error; Q1 = low-risk group; Q2 = middle-risk group; Q3 = high-risk group.

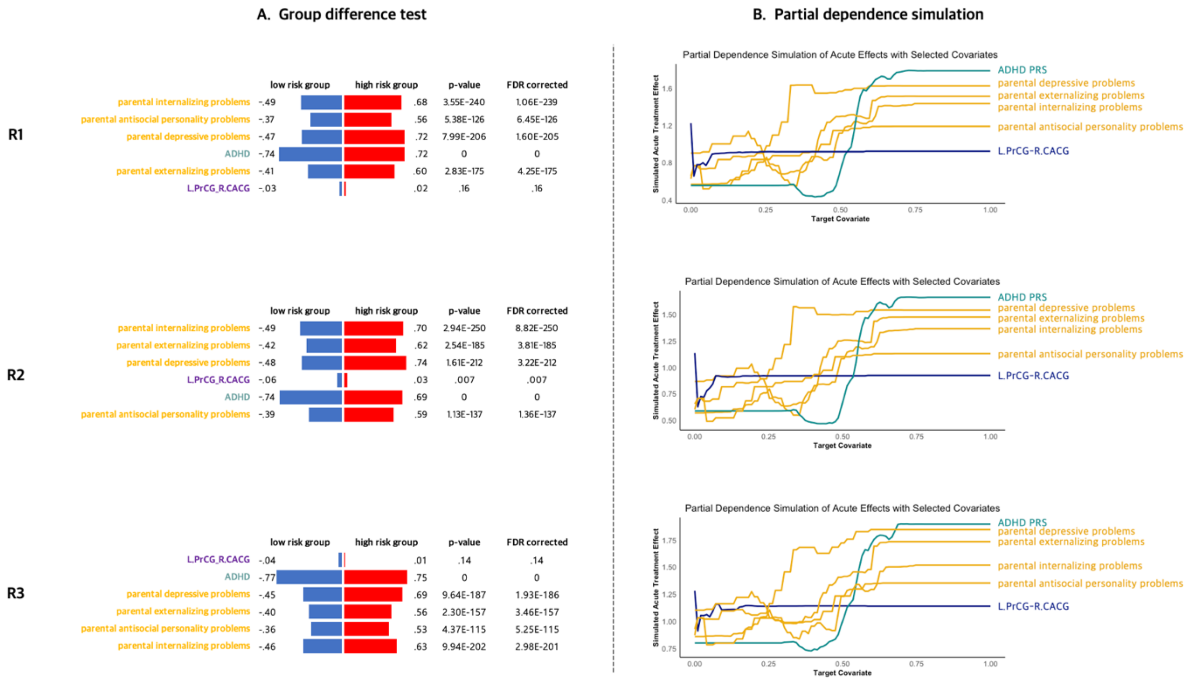


**Figure S4. The results of group difference test and partial dependence simulation at 2-year follow-up**

A. The comparison of three covariate values between low and high-risk groups across three Random Iterations. High-risk group significantly showed more parental mental health problems and higher polygenic scores of ADHD than low-risk group. The streamline count between left PrCG and right CACG was also stronger in high-risk group but showed inconsistent significancy across three Random Iterations. All covariate values are z-scaled.

B. The results of partial dependence simulation across three Random Iterations. All covariates showed increase in simulated treatment effect, except for the streamline count between left PrCG and right CACG that showed a slight initial increase but remained constant.

Yellow indicates environmental factors; green, polygenic risk scores; and blue, brain features.

R1 = random iteration 1; R2 = random iteration 2; R3 = random iteration 3; PRS = polygenic risk score; PrCG = left precentral gyrus; R.CACG = right caudal anterior cingulate gyrus.

**Table S6. The final model results of specificity analyses in the impact of stressful events on different mental disorders at 1-year and 2-year follow-ups**

| Outcome | ATE | p-value | FDR | Model fit | p-value | FDR | Heterogeneity Index | p-value | FDR |
|---|---|---|---|---|---|---|---|---|---|
| 1-year follow-up | | | | | | | | | |
| Random iteration 1 | | | | | | | | | |
| Depression | 1.00 | 1.11E-05 | 1.11E-05 | 1.06 | 9.16E-06 | 2.75E-05 | -1.77 | .78 | .78 |
| Anxiety disorder | .78 | 6.08E-04 | 6.13E-04 | 1.09 | 4.56E-04 | 6.76E-04 | 1.22 | .03 | .04 |
| Somatic problem | .73 | 3.11E-03 | 3.11E-03 | .85 | 7.20E-06 | 2.16E-05 | -1.16 | .71 | .71 |
| Conduct disorder | 1.45 | 7.30E-13 | 7.30E-13 | .97 | 1.31E-11 | 1.31E-11 | .87 | .05 | .05 |
| ODD | 1.33 | 8.45E-10 | 8.45E-10 | 1.02 | 9.83E-14 | 2.95E-13 | -1.06 | .74 | .74 |
| Random iteration 2 | | | | | | | | | |
| Depression | 1.05 | 3.67E-06 | 5.51E-06 | 1.08 | 2.68E-05 | 4.03E-05 | .01 | .50 | .78 |
| Anxiety disorder | .77 | 6.13E-04 | 6.13E-04 | 1.14 | 6.76E-04 | 6.76E-04 | 1.43 | .02 | .04 |
| Somatic problem | .75 | 2.09E-03 | 3.11E-03 | .91 | 5.66E-05 | 8.49E-05 | .16 | .45 | .68 |
| Conduct disorder | 1.48 | 1.17E-13 | 1.76E-13 | 1.00 | 1.39E-12 | 2.08E-12 | .93 | .03 | .05 |
| ODD | 1.36 | 1.74E-10 | 2.61E-10 | 1.01 | 3.93E-12 | 4.73E-12 | .24 | .43 | .64 |
| Random iteration 3 | | | | | | | | | |
| Depression | 1.04 | 3.66E-06 | 5.51E-06 | 1.04 | 4.57E-05 | 4.57E-05 | -.15 | .53 | .78 |
| Anxiety disorder | .79 | 4.31E-04 | 6.13E-04 | 1.12 | 3.40E-04 | 6.76E-04 | 1.33 | .06 | .06 |
| Somatic problem | .74 | 2.18E-03 | 3.11E-03 | .89 | 9.46E-05 | 9.46E-05 | .30 | .36 | .68 |
| Conduct disorder | 1.49 | 5.98E-14 | 1.76E-13 | 1.00 | 9.40E-13 | 2.08E-12 | .91 | .02 | .05 |
| ODD | 1.35 | 1.49E-10 | 2.61E-10 | 1.01 | 4.73E-12 | 4.73E-12 | .48 | .30 | .64 |
| 2-year follow-up | | | | | | | | | |
| Random iteration 1 | | | | | | | | | |
| Depression | 1.22 | 1.88E-06 | 2.82E-06 | .94 | 2.55E-09 | 4.29E-09 | 1.13 | .03 | .030 |
| Anxiety disorder | .68 | 5.20E-03 | .007 | .85 | 8.40E-04 | .002 | 1.11 | 3.99E-06 | 5.99E-06 |
| Somatic problem | .77 | 4.17E-03 | .006 | 1.15 | 1.66E-44 | 1.66E-44 | -21.68 | 1 | 1 |
| Conduct disorder | 1.67 | 5.29E-14 | 7.94E-14 | .92 | 7.66E-13 | 1.15E-12 | 1.25 | .04 | .066 |
| ODD | 1.40 | 6.81E-10 | 9.28E-10 | .97 | 3.65E-10 | 5.48E-10 | .78 | .004 | .004 |
| Random iteration 2 | | | | | | | | | |
| Depression | 1.19 | 3.17E-06 | 3.17E-06 | .90 | 4.29E-09 | 4.29E-09 | 1.40 | .03 | .030 |
| Anxiety disorder | .65 | .007 | .007 | .77 | .002 | .002 | 1.40 | 8.42E-06 | 8.42E-06 |
| Somatic problem | .73 | .006 | .006 | .93 | 8.42E-50 | 1.26E-49 | -21.49 | 1 | 1 |
| Conduct disorder | 1.66 | 8.46E-14 | 8.46E-14 | .92 | 5.68E-13 | 1.15E-12 | 1.09 | .07 | .066 |
| ODD | 1.39 | 9.28E-10 | 9.28E-10 | .93 | 6.34E-10 | 6.34E-10 | 1.05 | .002 | .004 |
| Random iteration 3 | | | | | | | | | |
| Depression | 1.26 | 7.36E-07 | 2.21E-06 | 1.02 | 2.88E-09 | 4.29E-09 | 1.29 | .01 | .030 |
| Anxiety disorder | .67 | .006 | .007 | .85 | .001 | .002 | 1.32 | 1.37E-06 | 4.11E-06 |
| Somatic problem | .77 | .004 | .006 | 1.08 | 6.42E-70 | 1.93E-69 | -20.05 | 1 | 1 |
| Conduct disorder | 1.73 | 6.53E-15 | 1.96E-14 | .95 | 3.31E-12 | 3.31E-12 | 1.20 | .06 | .066 |
| ODD | 1.45 | 1.33E-10 | 3.99E-10 | .94 | 3.41E-10 | 5.48E-10 | 1.12 | .003 | .004 |

Note. ATE = average treatment effect; ODD = oppositional defiant disorder. *** $p < .001$, ** $p < .01$, * $p < .05$

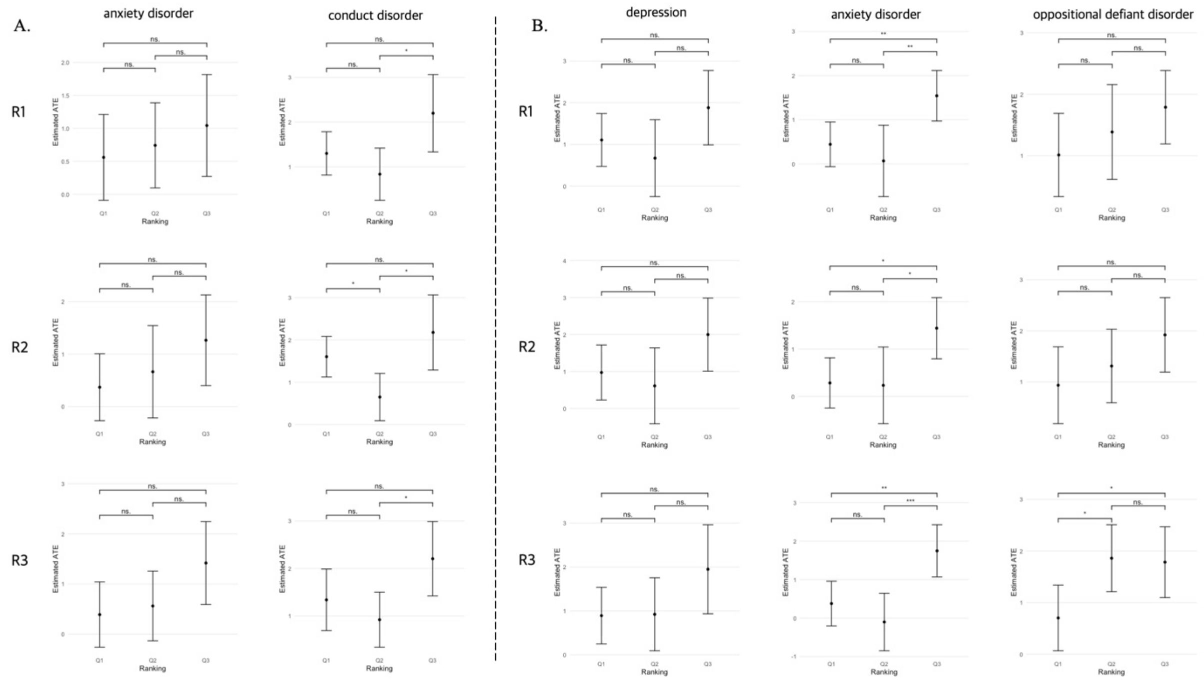


**Figure S5. The GATE test of specificity analyses in the impact of stressful events on different mental disorders at 1-year and 2-year follow-ups.**

A. The GATE test results of anxiety and conduct disorder at 1-year follow-up. To test the specificity of our results, the final model of ADHD at 1-year follow-up model was applied to other mental disorders (i.e., depression, anxiety disorder, conduct disorder, somatic problem, oppositional defiant disorder). The GATE test was utilized only for anxiety and conduct disorder that showed significant heterogeneity index (Table S6). All three Random Iterations demonstrated no significant individual differences on each disorder after experiencing stressful events.

B. The GATE test results of depression, anxiety disorder, and oppositional defiant disorder (ODD) at 2-year follow-up. Only the depression, anxiety, and ODD showed significant heterogeneity index (Table S6) and therefore the GATE test was applied. The results of all three Random Iterations suggested no significant individual differences on each disorder after experiencing stressful events.

ATE = average treatment effect; R1 = random iteration 1; R2 = random iteration 2; R3 = random iteration 3; ns. = not significant; Q1 = low-risk group; Q2 = middle-risk group; Q3 = high-risk group. *** $p < .001$, ** $p < .01$, * $p < .05$
